# Enhancing Fairness and Accuracy in Diagnosing Type 2 Diabetes in Young Population

**DOI:** 10.1101/2023.05.02.23289405

**Authors:** Tanmoy Sarkar Pias, Yiqi Su, Xuxin Tang, Haohui Wang, Shahriar Faghani, Danfeng (Daphne) Yao

**Affiliations:** Computer Science, Virginia Tech, Virginia, USA; Radiology, Mayo Clinic, Rochester, USA

**Keywords:** Ageism, Fairness, Diabetes, Machine Learning, Healthcare, BRFSS

## Abstract

While type 2 diabetes is predominantly found in the elderly population, recent publications indicate an increasing prevalence in the young adult population. Failing to diagnose it in the minority younger age group could have significant adverse effects on their health. Several previous works acknowledge the bias of machine learning models towards different gender and race groups and propose various approaches to mitigate it. However, those works failed to propose any effective methodologies to diagnose diabetes in the young population, which is the minority group in the diabetic population. This is the first paper where we mention digital ageism towards young adult population diagnosing diabetes. In this paper, we identify this deficiency in traditional machine learning models and propose an algorithm to mitigate the bias towards the young population when predicting diabetes. Deviating from the traditional concept of one-model-fits-all, we train customized machine-learning models for each age group. Our proposed solution consistently improves recall of diabetes class by 26% to 40% in the young age group (30-44). Moreover, our technique outperforms 7 commonly used whole-group sampling techniques such as random oversampling, SMOTE, and AdaSyns techniques by at least 36% in terms of diabetes recall in the young age group. We also analyze the feature importance to investigate the source of bias in the original model. We tested our approach on multiple datasets using multiple machine learning models and multiple sampling algorithms. Our code is publicly available at an anonymous repository - https://anonymous.4open.science/r/Diabetes-BRFSS-DP-C847

## I. Introduction

DIAGNOSING chronic diseases like diabetes is crucial, given the substantial global burden of diabetes-related complications and deaths, particularly in low- and middle-income countries [1], [2]. At the present rate of expansion, the International Diabetes Federation predicts that by 2045, a staggering 693 million individuals globally will be affected by diabetes [3]. The prevalence of diabetes, specifically type 2 diabetes, has been steadily increasing over the past few decades [4], [5]. While diabetes has traditionally been associated with the elderly population, recent studies indicate a rising prevalence of diabetes among the younger population as well [6]. According to CDC, by 2060, type 2 diabetes cases might increase by about 70-700% in the young population [7]. Young adults with diabetes face a significantly elevated risk of early health complications and even premature death compared to their counterparts without diabetes [8]. Younger people diagnosed with diabetes face an increased risk of developing early and severe complications, encompassing microvascular (retinopathy, neuropathy, ulceration, nephropathy) and macrovascular (cardiovascular, cerebrovascular, peripheral vascular) diseases [9]–[11]. Early detection and awareness can help the young population at risk take steps to prevent or delay type 2 diabetes, and early intervention can even reverse prediabetes [12], [13].

Machine learning (ML) has been increasingly integrated into the healthcare systems [14]–[16] because of its potential to assist clinicians and medical doctors in taking better care of patients. Many machine learning models have been applied to diagnose diabetes [5]. However, health data can be imbalanced, which could potentially lead machine learning models to learn patterns with existing bias from the provided data [17]–[22]. Data bias, if not addressed, can exacerbate and perpetuate inequalities in the performance of algorithms in different subgroups [23]–[25], particularly in historically underserved populations like female patients [26], black patients, or those with low socioeconomic status [27]. AI models can be susceptible to digital ageism, potentially leading to biased diagnoses that could harm patients [28], [29]. Moreover, AI models existing in the healthcare domain can show faithfulness issues [30]. It is of high importance that we evaluate the machine learning model before deployment ensuring social fairness [31], [32].

This paper identifies that the traditional machine learning models such as Logistic regression (LR), Multi-Layer Perceptron (MLP), Naive Bayes (NB), AdaBoost (AB), Random Forest (RF), and K Nearest Neighbor (KNN), trained on imbalance BRFSS (with only 15% representing the diabetes population) dataset [33], tend to misdiagnose diabetes more frequently in the younger population (30-44 years) compared to other subgroups. The recall of the positive class (Rec C1) in the 30-34 age group is only 30%, whereas the positive class (Rec C1) is 68-72% in the gender group and 66-84% in the ethnic group. Moreover, we bring attention to the fact that solely relying on AUROC can be misleading – while the overall group’s AUROC is 82%, the age group 30-34 exhibits a seemingly high 84% AUROC that masks the poor diabetes detection rate (recall C1) within that specific age group. So, we focus mostly on recall of class 1, balanced accuracy (average recall of class 1 and class 0), and AUROC for a fair comparison.

We propose an effective solution which successfully mitigates bias from young groups and increases type 2 diabetes (T2D) diagnosing sensitivity. None of the existing papers developed any effective machine-learning-based approach for effectively diagnosing T2D in the young population (30-44 years). To the best of our knowledge, our work is the first precision T2D diagnosis paper using machine learning and improving the diagnostic performance of machine learning models to diagnose T2D in young populations.

Our major contributions are:

1. We address a critical gap in the literature by developing a machine learning approach specifically designed for precision type 2 diabetes (T2D) diagnosis in a young adult population (30-44 years old). This is a new study to target this specific age group for T2D diagnosis using machine learning.
2. We show that multiple machine learning models and a number of sampling techniques (SMOTE, random sampling, etc.) fail to achieve fair performance in terms of detecting T2D in young adults.
3. We propose a bias correction technique specifically for improving T2D diagnosis in young adults. This demonstrates the effectiveness of subgroup-focused bias correction, promoting fairer and more accurate machine learning models in healthcare settings.

In this study, we train diverse machine learning models, incorporating a transparent model like logistic regression, to uncover the roots of bias and missed detections. This is achieved through a thorough analysis of association coefficients responsible for shaping model decisions.

## II Methods

### A. Dataset

The Behavioral Risk Factor Surveillance System (BRFSS) is an annual health survey conducted in the United States to monitor various health behaviors such as cardiovascular diseases, chronic diseases, diabetes, obesity, and other risk factors that contribute to the leading causes of death and disability [33]. The BRFSS datasets have been collected from all 50 states in the U.S. and the District of Columbia and included responses from over 400,000 participants which makes it one of the largest publicly available datasets related to public health. Each record contains an individual’s BRFSS survey responses on various health behaviors and risk factors such as tobacco use, physical activity, alcohol consumption, existing chronic diseases, and mental health. The survey also gathered demographic information such as age, gender, and race/ethnicity, which are helpful to explore important correlations and even causation.

This survey is conducted every year, and the CDC makes it publicly available for research. In this study, we selected the BRFSS dataset from 2021, which contains more than 400,000 subject information and 330+ attributes from each subject. The BRFSS dataset is a valuable resource for identifying health disparities and evaluating the effectiveness of public health programs and policies. However, the dataset is not free from challenges because a large portion of attribute values are missing (25%) and this dataset can be a highly imbalanced data imbalance (diabetes class 15%).

### B. Data Preprocessing

According to the literature on the risk factors for diabetes, [5], [34], we selected 30 attributes including diabetes labels. Subjects aged over 30 are selected for this study [5], [35], [36]. We create age groups spanning 5 years starting from 30 up to 80+ years. The selected cohort contains 200,136 subjects information where 169,296 subjects (84.6%) are diabetes negative and 30,840 subjects (15.4%) are positive.

As we mentioned before, this dataset is not balanced for the age, gender, and racial subgroups. In the age group 30-34, the number of negative cases is 10,864 but the number of positive cases is only 324. The age groups 35-39 and 40-44 are also highly imbalanced as the positive-negative ratio is only 4.4% and 7.2%.

The selected variables fall into three distinct types: nominal, ordinal, and binary. Nominal variables lack any inherent order, ordinal variables possess a meaningful order, and binary variables exclusively hold two distinct values. For example, the presence of high blood pressure, cholesterol, or heart disease can be represented by a binary variable. BMI category, education level, and income level are considered ordinal variables. On the other hand, marital status and race are nominal variables. Binary variables are represented using a binary encoding, where 1 signifies a positive outcome and 0 represents a negative outcome. Ordinal variables are encoded using integers, preserving their meaningful order. Nominal variables are transformed into one-hot encoding for appropriate representation. The selected variables are listed in Fig. 6(c) .

### C. Bias Mitigation Approach

We utilized a modified and enhanced version of the prioritized (DP) bias correction method (Fig. 1) which is inspired by [37] prioritizes a specific subgroup, the young age group in this case, that suffers from data imbalance. We incrementally replicate data points of the minority class (diabetes positive indicated by class 1) and choose an optimal unit of replication based on the model performance. As a result, the enriched training set contains the original samples as well as the replicated samples. However, the vanilla DP technique by [37] has several shortcomings, including the selection of replication units.

**Fig. 1.**
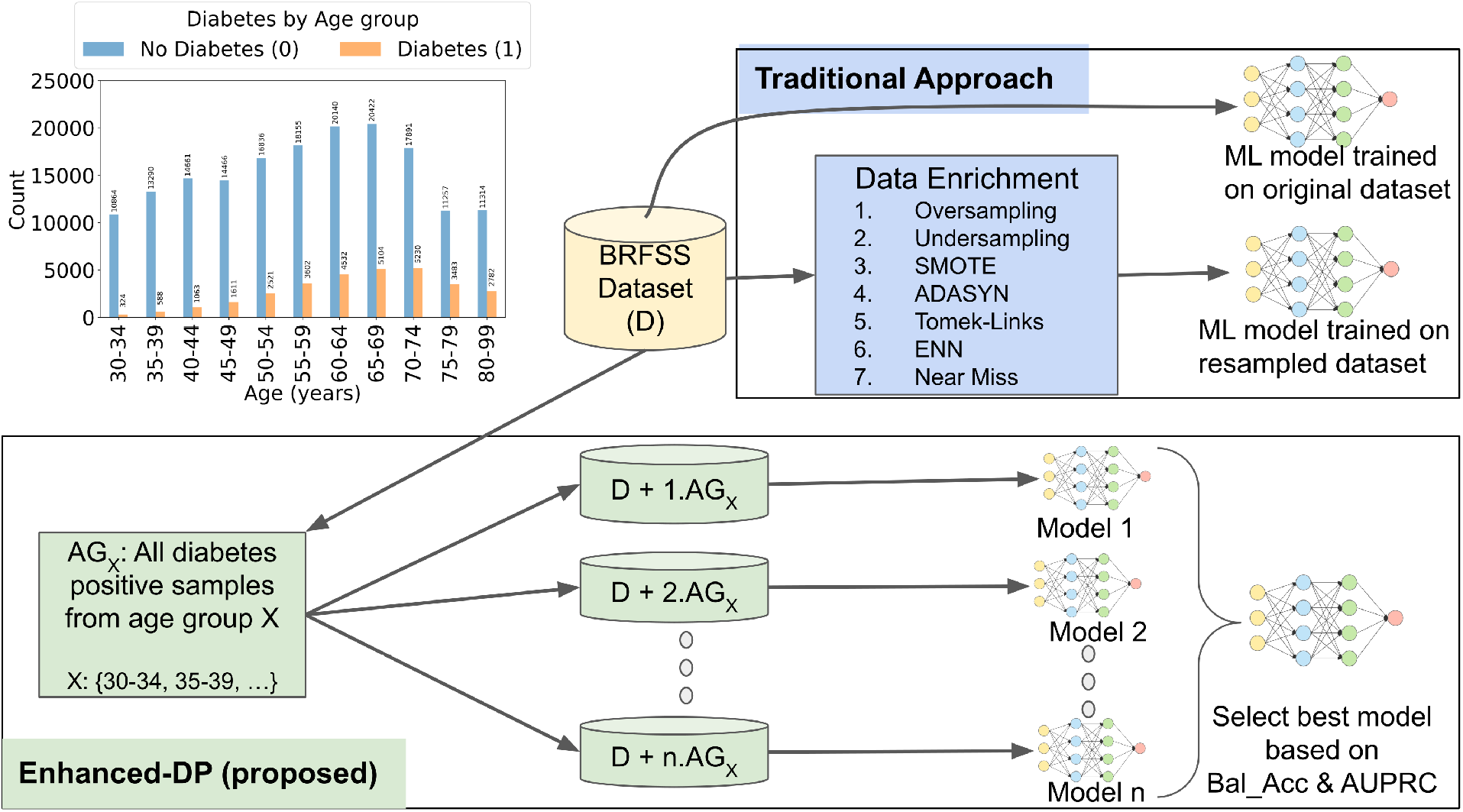
The Enhanced-DP approach in contrast to traditional approaches, enriches the minority age groups and creates new training sets by replicating diabetic samples (1 to ***n*** times) from a minority age group. ***n*** machine learning models are trained on each of the ***n*** versions of the training sets. The best model is selected based on performance metric balance accuracy (Bal Acc) and area under precision and recall curve (AUPRC). The top left bar chart represents age distribution (histogram) in the original dataset.

Our proposed Enhanced-DP technique replicates all samples of the diabetes class from the young population up to *n* times.

Each time the duplicated sample units are merged with the original dataset, including the original set. In our experiment, we set the maximum DP unit “n” based on the lowest subgroup positive-negative sample ratio. In order to achieve a balanced set, the lowest subgroup ratio is for the age group 30-34. The number of negative cases is 10,864 but the number of positive cases is 324. So, the ratio is 34, which is the limit of DP unit *n*. We also employed early stopping to select the best *n* quickly as the performance curve contains a global maximum point. This makes our Enhanced-DP algorithm faster compared to the original DP algorithm [37].

Each training set containing the original set and the duplicated *n* 1 to 34) units is used to train a single model. In this way, we train 35 models (34 models trained on the enriched training sets and 1 model trained on the original training set) and select the best-performing model based on balanced accuracy and the area under the curve (AUC) of the minority class (C1) precision-recall curve, denoted by PRC C1. We identify the top three machine learning models with the highest balanced accuracy values and select the model that gives the highest PR C1 for that particular age group. This selected model is used only for diagnosing diabetes for that particular group, e.g., the Enhanced-DP model trained with duplicated age-group 30-34 samples is used to diagnose new patients from age-group 30-34 years.

### D. Sampling Algorithms

A frequently employed technique for addressing the challenges of data imbalance is the utilization of sampling methods. To evaluate the effectiveness of the sampling algorithms detecting diabetes in young groups, we first compared multiple sampling algorithms, including (1) random oversampling, randomly duplicating instances from the minority class [38]; (2) random undersampling, randomly removing instances from the majority class [38]; (3) SMOTE (Synthetic Minority Over-sampling Technique), creating synthetic samples for the minority class by interpolating between existing instances [39]; (4) ADASYN (Adaptive Synthetic Sampling), similar to SMOTE, but generating more synthetic samples for difficult-to-learn instances [40]; (5) Tomek Links, removing instances from the majority class that are close to instances in the minority class [41]; (6) ENN (Edited Nearest Neighbors), removing instances from the majority class that are misclassified based on the nearest neighbors from both classes [42]; and (7) NearMiss, selecting instances from the majority class based on their distance to instances in the minority class [43]. These sampling techniques have been proven to be effective in the literature, however, these techniques are not tailored to tackle subgroup biases. As a result, diabetes is misdiagnosed in the young population. Therefore, we utilize a new concept of using one model for a single group which deviates from the traditional one model-fits-all ideology.

### E. Machine Learning Models

We selected six commonly used ML models including Logistic Regression (LR) [44], Random Forest (RF) [45], Adaboost (AB), Multilayer Perceptron (MLP) [46], and Naive Bayes (NB) to evaluate the effectiveness of the sampling strategy. We purposely selected simple models such as logistic regression because of their interpretability. It is very important for the science community, especially for healthcare to create nterpretable models to find out the root cause of a prediction, however, [47] evaluated 511 scientific papers across different ML domains and identified a notable deficiency in reproducibility metrics, including dataset and code accessibility in clinical ML domain papers. Moreover, complex and high-end models which have recently been applied to the healthcare domain might pose difficulties in reproducibility [48]. Using simple models like logistic regression will pave the way both for explaining the results and making it easy for other researchers to reproduce the model and the same results.

In the RF, we selected 100 trees or estimators to achieve optimal performance. In our MLP configuration, we used a single hidden layer with 100 ReLU-activated neurons and optimized training with the Adam gradient descent optimizer for efficiency and effectiveness. For Naive bayes we utilized the Gaussian kernel. For Adaboost we select the default settings from scikit-learn Python library. We repeated the training process at least five times with different test train splits to ensure consistent performance and calculate the standard deviation of the performance.

For LR, we utilized the liblinear equation to establish the decision boundary between positive and negative classes. This analysis can provide valuable insights into feature importance, offering a comprehensive understanding of how each variable contributes to the model’s outcome. We investigate the features responsible for biased outcomes from the original model. The logistic regression estimates the probability from the coefficients and the corresponding feature value which makes it a white box and fully interpretable.

To examine the potential unfairness in the datasets, we calculated class-based accuracy, recall, AUROC, and AU-PRC.

Nonetheless, our primary focus lies in prioritizing the recall of the positive class, also known as sensitivity, as the detection of diabetes holds paramount importance in our specific case. The dataset undergoes a random division into three disjoint sets namely training (60% or 120,081 samples), validation (20% or 40,027 samples), and testing set (20% or 40,028 samples) through the utilization of the Python sklearn library. The training dataset serves as the foundation for model training, while the testing dataset remains consistent across all the experiments, ensuring the robustness and comparability of our results. Only the training set is used for DP, ensuring no data leakage. The performance of the models in each experiment is recorded as the average value of 3-5 independent trials.

### F. Model Calibration and Threshold Tuning

Before mapping probabilities into labels, we calibrate the predicted probabilities for each model on the validation set. Calibration involves adjusting the distribution of probabilities to enhance accuracy. We calibrate the output probabilities using the Isotonic Regression [49] technique. We then perform threshold tuning to find the optimal threshold based on balanced accuracy and the F1 C1 score. Threshold tuning is the process of selecting an optimal threshold value in binary classification models. In these models, predictions are typically expressed as probabilities, and a threshold is applied to determine the class label. Adjusting this threshold can impact the trade-off between false positives and false negatives.

At first, we identify the top 3 thresholds that produce the highest F1 C1 scores and then select the optimal threshold that generates the highest balanced accuracy for all samples in the validation dataset. For some subgroups, only a few samples (*<* 100) are present in the validation set. Selecting the threshold based on subgroup samples may overfit the validation set. So, we select a threshold based on the whole validation performance. We calculate the optimal threshold to be 0.195 from 10 independent trials by averaging the best thresholds from each trial. We use the F1 score and balanced accuracy jointly to select a model with a balanced performance.

## III. Results

### A. Performance of Original Model

The machine learning models trained on the original training set show different performances for the minority diabetes-positive group (C1) and the majority-negative group (C0). Fig. 2 (a) and (b) shows the performance of the original logistic regression model on 2021 and 2015 datasets. The whole group recall C1 is 0.70-0.72 whereas recall C0 is 0.77-0.78. For males, recall C1 is 0.72 and for Female recall C1 is 0.68. The ethnic groups such as White, Asian, and multicultural show recall C1 of 0.66, 0.66, and 0.73 respectively. On the other hand, Black, Indian, and Hawaiian show recall C1 of 0.84, 0.82, and 0.84 respectively.

**Fig. 2.**
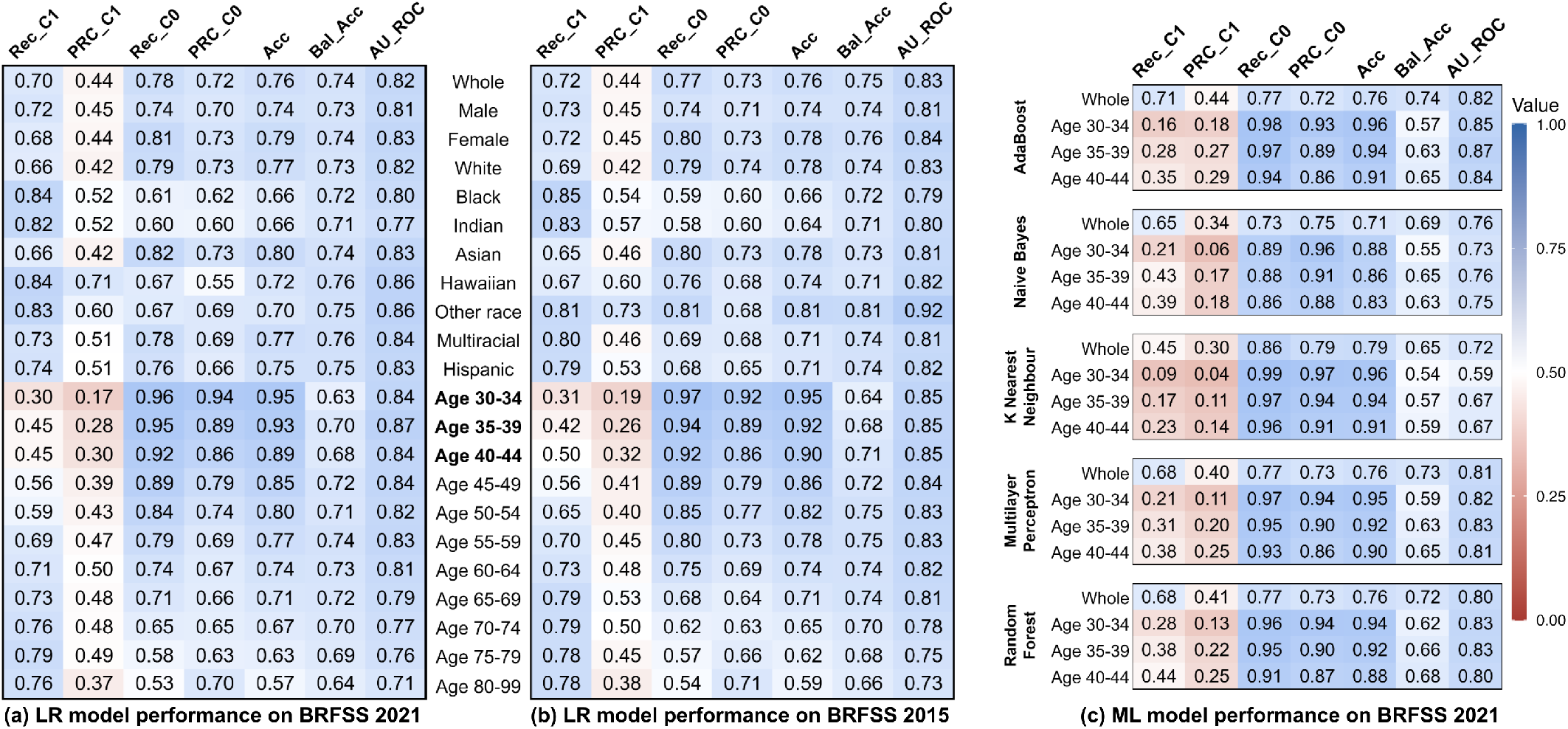
Performance of logistic regression model trained and tested on the original (a) BRFSS 2021 and (b) BRFSS 2015. (c) Performance of multiple machine learning models for the young adult age group (30-44 years) along with the whole group. The x-axis represents performance metrics where Rec, PRC, Acc, Bal Acc, AUROC, represent Recall, Area Under the Precision-Recall Curve, Accuracy, Balanced Accuracy, and area under the ROC curve respectively. C1 and C0 stand for class 1 (diabetes positive) and class 0 (diabetes negative) respectively. The Y-axis represents the different subgroups.

However, the young age group suffers the most from missed detection of diabetes. For age groups [30-34], [35-39], and [40-44] recall C1 is only 0.30-0.31, 0.42-0.45, and 0.45-0.50 respectively. This means out of 100 diabetic patients aged 30-34, 70 patients are misdiagnosed. The Adaboost, Naive bias, random forest, and KNN models also show similar performance, represented in Fig. 2 (c). The standard deviation of each metric is less than 0.04 (from multiple experimental trials) unless mentioned otherwise. The model behaves similarly in the BRFSS 2015 dataset (this is not shown due to spaceconstraints).

In imbalanced datasets, commonly employed metrics like AUC ROC and accuracy can be misleading and do not accurately represent the performance of the minority class. Despite potentially poor performance in the minority class, these metrics might indicate falsely elevated values. The AUROC of the whole population, age groups [30-34], [35-39], and [40-

44] are 0.82, 0.84, 0.87, and 0.84. However, these age groups show very poor recall C1 which is not reflected with AUROC. In contrast to the AUROC and accuracy metrics which can be overly optimistic, we use Recall C1 to measure the true detection rate reflecting the performance of the model.

### B. Enhanced-DP model improves diagnostic accuracy

The Enhanced-DP model for three age groups improves the diabetes detection accuracy significantly (Fig. 3 (a)). Fig. 3 (b) shows the performance difference between the Enhanced-DP model and the original model in terms of positive recall and balanced accuracy. Diff Rec C1 means subtracting the recall of class 1 of the original model from the Enhanced-DP model. The Enhanced-DP model for age groups [30-34], [35-39], and [40-44] improves the positive recall by 41% (SD 6.4%), 32% (SD 6.4%), and 24% (SD 2.8%) respectively. This means the Enhanced-DP models are better at detecting diabetes in the young population.

**Fig. 3.**
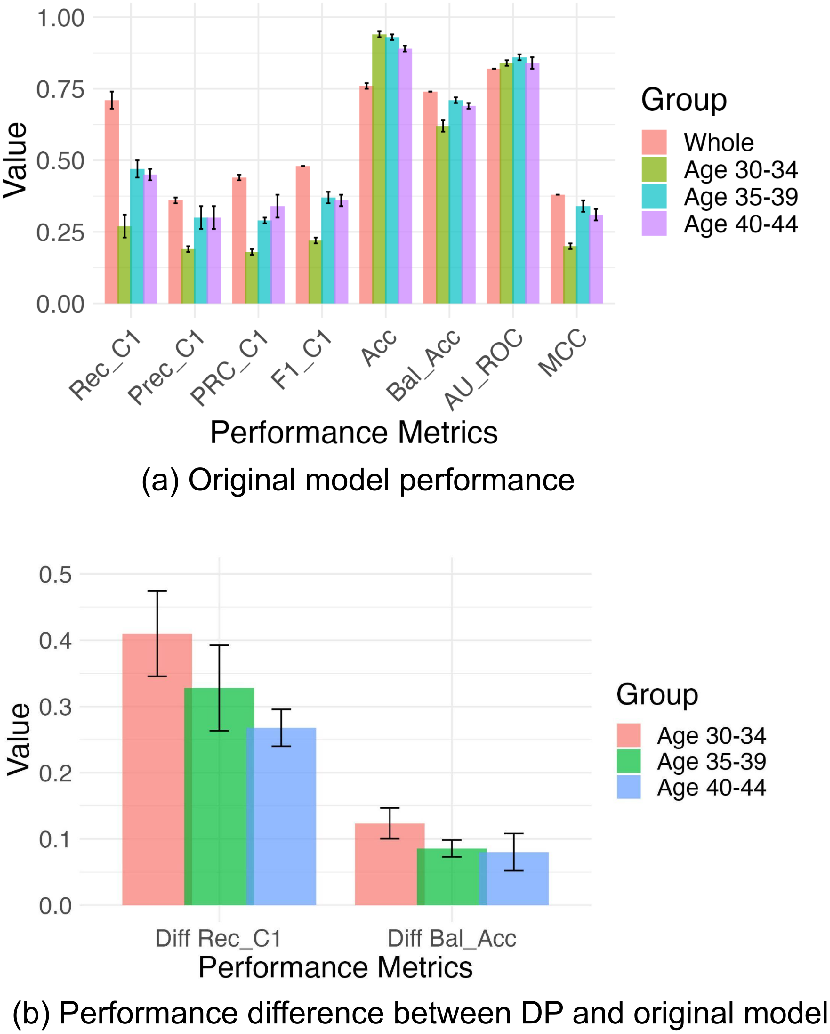
Performance of the original logistic regression model (a) when tested on the whole population and minority age group. (b) Performance difference between Enhanced-DP and original model Logistic Regression model. “Diff Rec C1” means subtracting the recall of class 1 of the original model from the Enhanced-DP model and “Diff Bal Acc” means subtracting the balanced accuracy of the original model from the Enhanced-DP model. Positive values indicate performance improvement from the original model. The error bars represent the standard deviation of the experiment results.

**Fig. 4.**
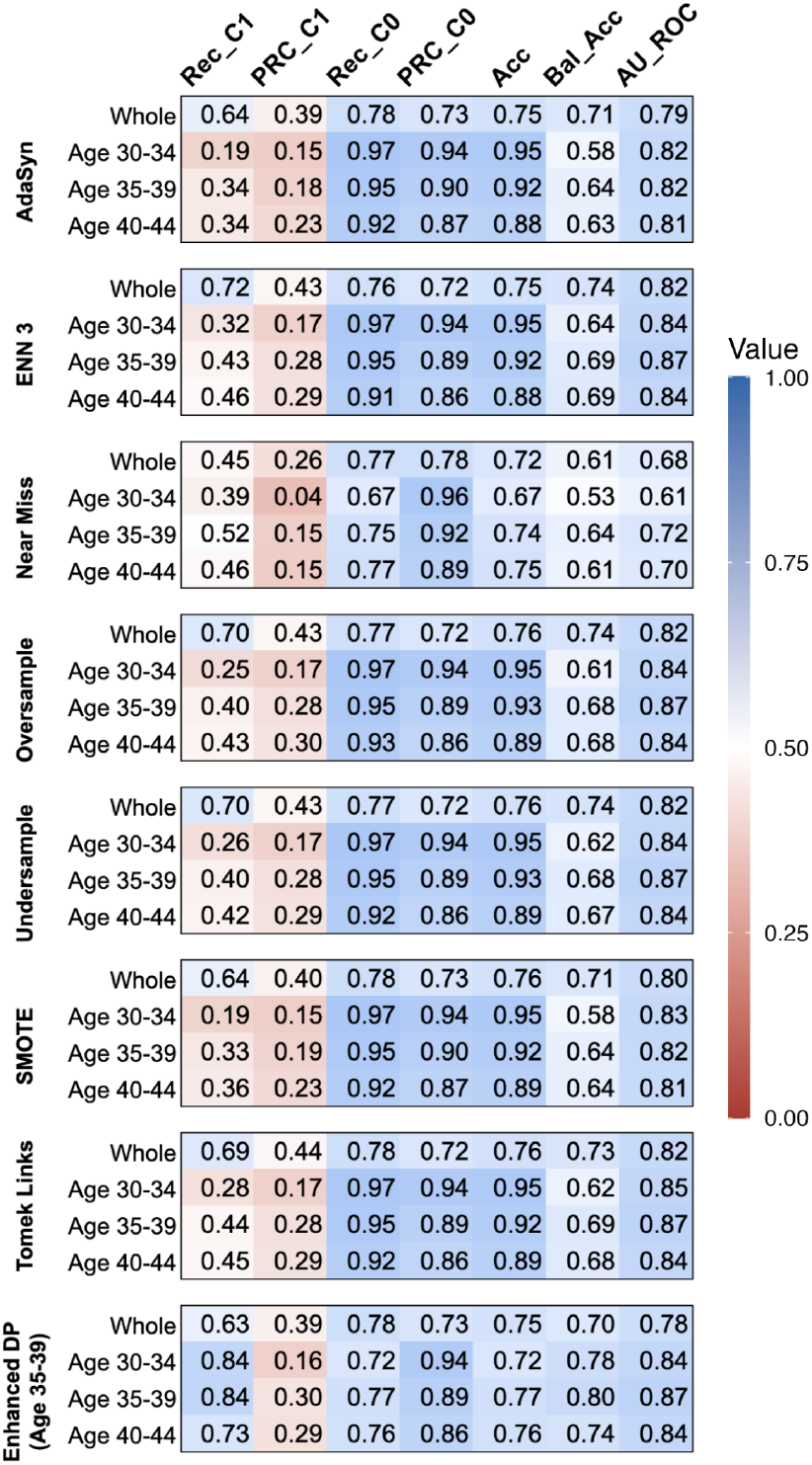
Comparing whole group sampling method performance with Enhanced-DP (age group 35-39) for the young adult age groups 30-34, 35-39, and 40-44 along with the whole group.

On the other hand, Diff Bal Acc means subtracting the balanced accuracy of the original model from the Enhanced-DP model. The balanced accuracy (average recall of both C0 and C1 classes) is also improved by 13% (SD 2.3%), 10.5% (SD 1%), and 7.7% (SD 2.8%) for the Enhanced-DP model.

### C. Whole-population sampling

The whole group-based sampling approach doesn’t improve the detection rate in the young group Fig. III-C. Moreover, AdaSyn (Rec C1 0.19), SMOTE (Rec C1 0.19), Tomek-Link (Rec C1 0.28), Random oversampling (Rec C1 0.25) and undersampling (Rec C1 0.26) methods decrease the original detection accuracy (Rec C1 0.30) for age group [30-34]. A similar performance decrease is observed in the other two age groups [35-39] and [40-44]. Near Miss and ENN merely improve the recall C1 by 9% and 2%. On the other hand, the respective Enhanced-DP models (i.e. trained on age group [30-34] and applied to age group [30-34]) improve the positive recall by at least 24%.

The whole group sampling methods also show poor performance in terms of balanced accuracy. For example, for the age group [35-39] all of the methods decrease the balanced accuracy from the original value. On the other hand, the Enhanced-DP model increases balanced accuracy by 3-9%.

### D. Segmented training

We compared the performance of models trained on age-segmented datasets, where the original dataset was divided into groups with 5-year and 15-year spans. Each model was trained and tested on data from the same or overlapping age groups. For example, a model trained on individuals aged 30-34 was evaluated using a test set from the same age range. Similarly, a model trained on the 30-44 age group was tested across the 30-34, 35-39, and 40-44 age groups. Each model was calibrated using the Isotonic Regression and model-specific decision threshold was calculated. We applied all seven resampling techniques outlined in Fig. III-C and reported the best performance for each model. As shown in Fig. 5, the DP method consistently performed the best. Interestingly, models trained on 15-year spans outperformed the baseline model trained on the entire dataset. These findings were also consistent with results obtained from the BRFSS’15 data.

**Fig. 5.**
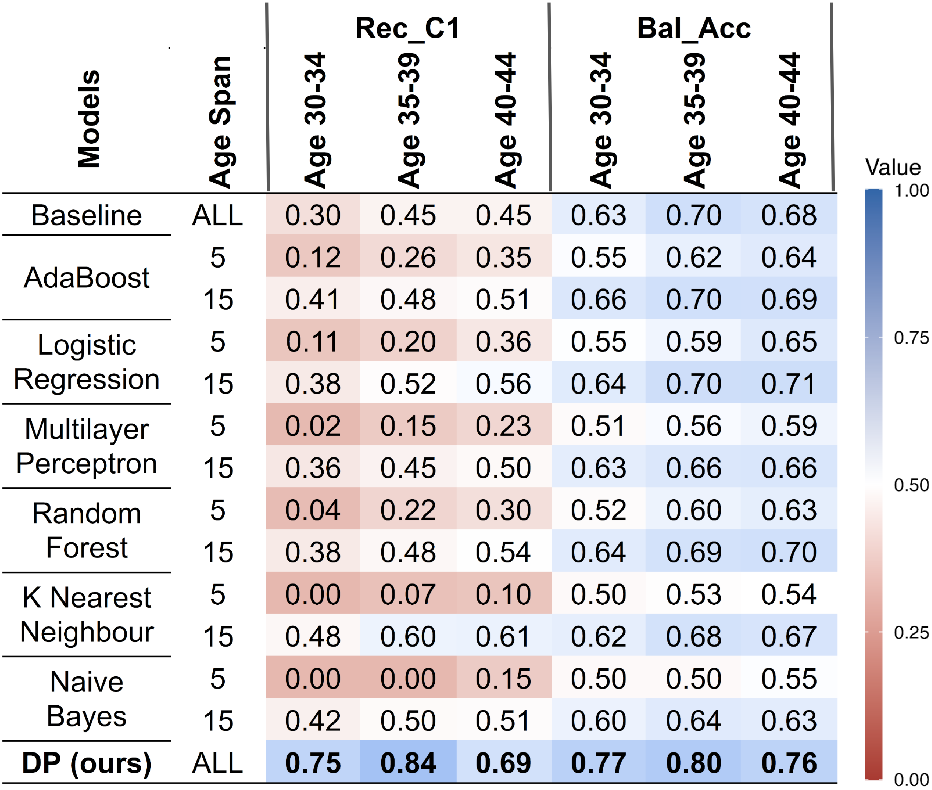
Models trained on age-segmented training sets composed of age groups spanning 5 years or 15 years. The models are trained and tested on the same or overlapping age group. We utilized all 7 resampling techniques (Fig. III-C) and reported the best performance for each model. The baseline model represents logistic regression model trained on the whole training set whereas DP is our proposed model.

### E. Cross-group performance

Enhanced-DP model trained for a specific age group is applied to all other age groups Fig. 6 (a) and (b). For example, a DP model trained for age group [30-34], is tested on age groups [30-34], [35-39], and [40-44]. We also compare the whole group’s performance with the DP models. The original model shows very poor recall C1 for all three age groups. Interestingly, the DP model trained for the age group [30-34] can also be applied to age groups [35-39] and [40-44]. DP model for age group [35-39] shows the highest recall C1 of 73% - 84% for all age groups. The balanced accuracy is also high when one DP model is applied to another age group.

**Fig. 6.**
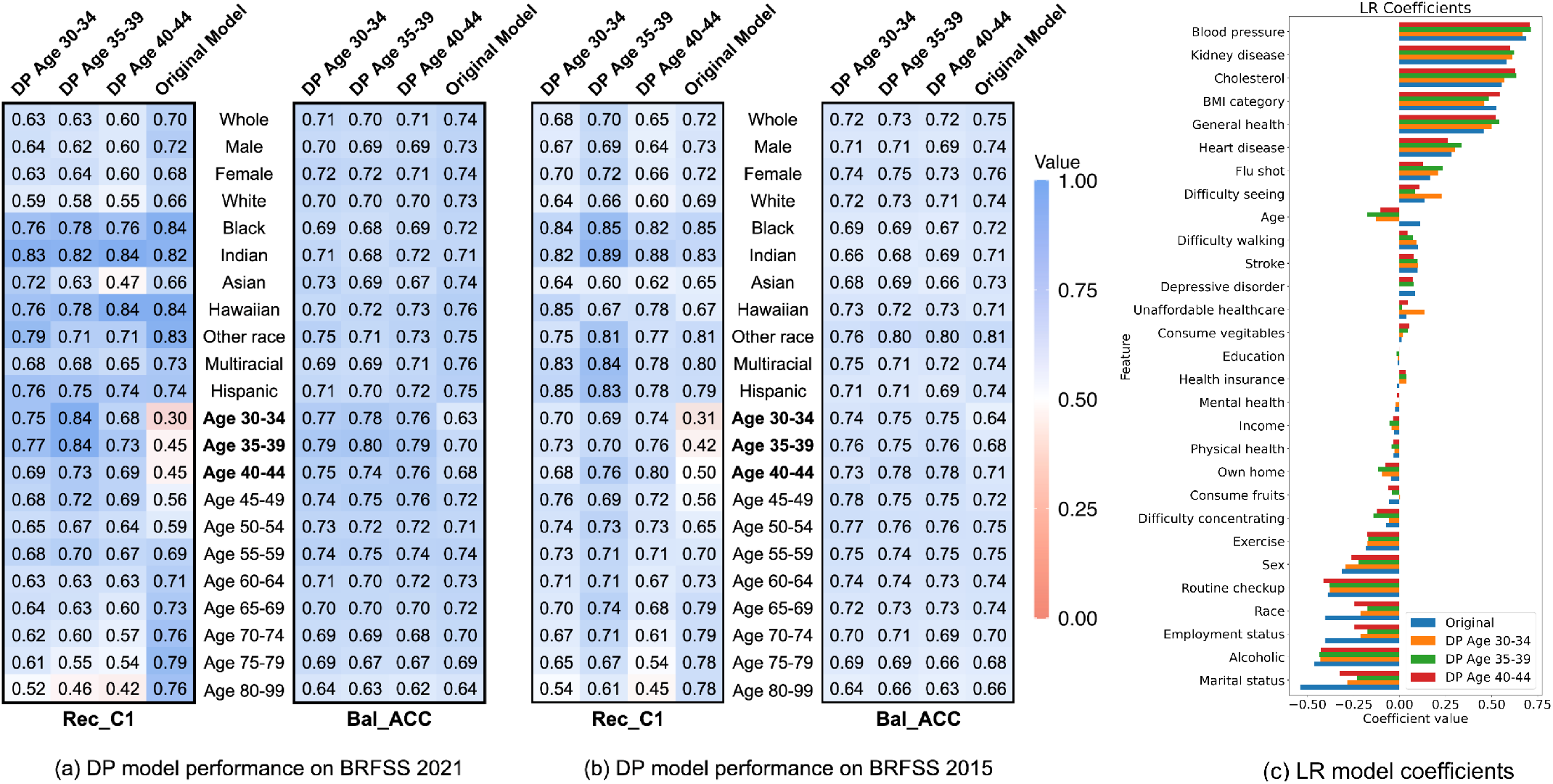
Cross-group performance analysis using class 1 recall (Rec C1) and balanced accuracy (Bal Acc) on (a) BRFSS 2021 and (b) BRFSS 2015. In each subfigure, each column corresponds to an Enhanced-DP model trained for a specific subgroup. Each row represents a subgroup that a model is evaluated on. (c) Represents logistic regression model coefficient values which are associated with each feature from the original model and Enhanced-DP models. The one hot encoded feature (marital status, employment status, and race) coefficients are averaged

On the other hand, the DP model trained for the young age group shows poor performance when applied to gender or ethnic groups for which the DP model is not trained. The recall of class 1 goes down by 8% and 19% when the DP model for the age group [40-44] is applied to the female and Asian groups respectively. The balanced accuracy also declines if age group DP models are applied to female or Asian groups.

### F. Feature analysis

One of the major motivations for selecting a logistic regression model is its interpretability. [50] showed that deep learning models are mostly black-box and cannot always be correctly interpretable.

Fig. 6 (c) shows the logistic regression model coefficients of the original and the Enhanced-DP models. It shows that the original model had a strong correlation with the subject’s race (-0.40), employment status (-0.40), and marital status (-0.54). The Enhanced-DP model reduces the strength of these attributes by at least 19%. Moreover, age was positively correlated in the original model while it was negatively correlated in the Enhanced-DP models. Other attribute coefficients show minor changes.

## IV Discussion

The machine learning trained on the original dataset shows poor diabetes diagnosis performance in the young group.

Because the diabetes data is limited in the original dataset the traditional machine learning model tends to pick the general statistics to build a diagnostic model. As a result, the models show poor performance in the minor young group. Whole group-based data enrichment such as sampling methods cannot overcome the problem of poor performance as it doesn’t enhance the young group. Some of them such as SMOTE, a popular sampling method which is well known for removing bias from imbalanced datasets, decrease the performance. This is because these sampling methods are not equipped to reduce disparate ratios in the minority group. Moreover, we tested multiple and diverse machine learning models. However, all the models show consistently poor performance in this scenario.

Finally, we proposed an enhanced version of the double prioritized bias correction technique (DP) to make the model effective and useful for the young age group. By replicating the minor group population dynamically, the technique improves the model’s performance. However, one DP model can be targeted for one particular group. The results show that the DP model trained for the young age group is not applicable for both gender or ethnic groups. This limitation can be easily overcome by using multiple DP models for each minor group. However, one of the limitations of this approach is the use of multiple models compared to a single model. This limitation only applies in the training phase but not in the application phase. Training multiple DP models will take more time than a single model approach but the DP model will take the same time during diagnosis as only one model is going to be used for predicting a particular sample. Interestingly, this technique has broader applications beyond the specific dataset evaluated and can be utilized in any domain to overcome bias or data limitation challenges.

We also investigated the root cause of bias in the original model by visualizing the coefficients of the logistic regression model. Being a white box model, we can easily understand the feature importance and the correlation of each feature with the detection probability. The original model shows a strong correlation with non-medical attributes such as marital status, employment status, and race. The DP model decreases the strength of correlation with these factors significantly. Moreover, the original model is positively correlated with age. It means the diabetes-positive probability increases with age. However, this positive correlation was one of the key factors why the original shows poor performance in the young age group.

We can also use real-world data in the form of the observational medical outcomes partnership (OMOP) common data model (CDM) or any other data sources. The core methodology will be the same as the DP method is model-agnostic as well as data-type agnostic. The OMOP-CDM might need to be preprocessed (e.g., relevant cohort selection, data cleaning, data scaling) before feeding to our proposed DP model. Our approach can be applied to any kind of data including OMOP-CDM.

In conclusion, we identified a major deficiency in the traditional machine learning and commonly used sampling approach when it comes to diagnosing diabetes in the young age group which is the minority population in this case. We proposed an enhanced DP method to overcome this issue and improved diagnostic performance significantly. This approach has the potential to mitigate bias issues in the diagnosis of diabetes among young adults, offering a pathway toward more equitable and accurate healthcare practices in this demographic.

## Data Availability

All data produced are publicly available online at www.cdc.gov

https://www.cdc.gov/brfss/annual_data/annual_data.htm

